# Clinical heterogeneity based on race and sex within a large cohort of inclusion body myositis patients

**DOI:** 10.1101/2022.05.24.22275537

**Authors:** E. Harlan Michelle, Iago Pinal-Fernandez, Maria Casal-Dominguez, Jemima Albayda, Julie J. Paik, Eleni Tiniakou, Brittany Adler, Christopher A. Mecoli, Sonye K. Danoff, Lisa Christopher-Stine, Andrew L Mammen, Thomas E. Lloyd

**Affiliations:** Department of Neurology, Johns Hopkins University School of Medicine, Baltimore, Maryland; Muscle Disease Unit, Laboratory of Muscle Stem Cells and Gene Regulation, National Institute of Arthritis and Musculoskeletal and Skin Diseases, National Institutes of Health, Bethesda, Maryland; Faculty of Health Sciences and Faculty of Computer Science, Multimedia and Telecommunications, Universitat Oberta de Catalunya, Barcelona, Spain; Department of Medicine, Johns Hopkins University School of Medicine, Baltimore, Maryland

## Abstract

**Background and Objectives:** Sporadic inclusion body myositis (IBM) is the most common acquired myopathy in individuals over age 50. The disorder is slowly progressive and while many therapies have been investigated, response has generally been poor. Clinical heterogeneity may influence treatment responsiveness; however, data regarding heterogeneity in IBM is limited and often conflicting. We aim to identify clinically distinct subgroups within a large IBM cohort, as well as prognostic factors for disease progression.

**Methods:** Clinical, histologic, radiologic, and electrophysiologic data were analyzed for all patients with IBM and other forms of myositis enrolled in a longitudinal cohort from The Johns Hopkins Myositis Center from 2003-2018. Univariate, multivariate, and graphical analyses were used to identify prognostic factors in IBM patients.

**Results:** Among the 335 IBM patients meeting inclusion criteria, 64% were male with an average age of disease onset of 58.7 years and a delay to diagnosis of 5.2 years. Initial misdiagnosis (52%) and immunosuppressant treatment (42%) were common. Less than half (43%) of muscle biopsies demonstrated all three pathologic hallmarks: endomysial inflammation, mononuclear cell invasion, and rimmed vacuoles. Black patients had significantly weaker arm abductors, hip flexors, and knee flexors compared to non-Black patients but were less likely to develop dysphagia. Female patients had stronger finger flexors and knee extensors compared to their male counterparts but were more likely to develop dysphagia. A significant number (20%) of patients had an age of onset less than 50 years. This group of younger patients was weaker at their first visit; however, this may be accounted for by a longer disease duration at first visit.

**Discussion:** Although IBM has long been considered a disorder predominately of older, White men, female, and non-White patients comprise a significant proportion of the IBM population. Our study demonstrates that female and Black patients have distinct clinical phenotypes within the overarching IBM clinical phenotype.

## INTRODUCTION

Sporadic inclusion body myositis (IBM) is the most common acquired myopathy among individuals older than 50 years of age. Gradual progression of muscle weakness and atrophy, particularly of the finger flexors and quadriceps muscles, and muscle biopsies showing endomysial inflammation with auto-invasive T-cells and rimmed vacuoles are hallmarks of the disease. Although there is evidence for dysregulation of the immune system in IBM, multiple trials of immunosuppressive agents have failed to show a benefit.^1–5^ Both degenerative and neoplastic processes have been suggested as alternative mechanisms.^6,7^

Of the relatively few studies that have closely examined the clinical symptoms and course of IBM, most have investigated small cohorts (mean 47 patients) with limited follow-up (mean 2.3 years).^8–16^ Although these studies have described an overall consistent IBM phenotype, there is evidence of substantial variability both within and between studies as regards presentation, clinical progression, and accumulation of debility. While the majority of patients in these studies had an age of onset in their late fifties, a considerable number (5-20%) developed symptoms at less than 50 years of age. Onset of weakness in the lower extremities was most common, however onset in the upper extremities was reported in 10-20% of patients and onset in the bulbar muscles in 5-10% of patients. About half of patients overall developed dysphagia during their disease course, and there was considerable variability in the progression of weakness in other skeletal muscles both between muscle group and individual patients Most patients eventually required the use of an assistive device for ambulation, however there was a wide range in the time from diagnosis to the need for an assistive device (range 6-16 years). Data on mortality was mixed with some studies showing increased mortality in IBM patients, often related to respiratory causes,^16–18^ whereas others demonstrated no impact of IBM on mortality.^11,12^

Data on prognostic factors has likewise been limited and often conflicting. Several studies have shown a more rapid functional decline in patients with older age of onset.^9– 11,14^ However, others found mixed results^13,19^ or no correlation between age of onset and progression of weakness,^12^ and one study reported a more rapid decline in pinch strength among individuals with a younger age of onset.^15^ Oldroyd et al reported a more rapid decline in pinch and knee extensor strength in men, however, sex was not found to have an association with strength decline in other studies.^12,13^ The presence of anti-NT5c1A autoantibodies has also been reported as a poor prognostic factor in some studies,^15,20^ although in others, no correlation was seen between seropositivity and prognosis.^21,22^

These studies suggest that IBM is a much more heterogeneous condition than has previously been considered. A larger study of IBM is important to identify distinct subgroups of IBM patients and the prognostic factors that may influence their clinical trajectory. Such information can better inform patients about their expected disease course and can influence the design of clinical trials. We conducted a prospective longitudinal cohort study of 335 IBM patients followed at the Johns Hopkins (JH) Myositis Center to identify IBM subgroups and their associated prognostic indicators.

## METHODS

### Patients

All patients who provided informed consent to be enrolled in the JH Myositis Center Registry between 2003 and 2018 with a diagnosis of sporadic IBM were reviewed. All patients were evaluated by a JH Myositis Center physician. Those meeting at least one of the following criteria were included for analysis: Griggs possible, European Neuromuscular Centre (ENMC) 2011 probable, or Lloyd-Greenberg data-derived criteria (DDC) for IBM.^23–25^ We included as comparators all those myositis patients that were positive for autoantibodies recognizing Mi-2, NXP-2, TIF1-γ, MDA-5, Jo-1, PL-7, PL-12, SRP, or HMGCR by at least two different techniques from among the following: ELISA, immunoprecipitation of *in vitro* transcribed and translated protein, line blotting (EUROLINE myositis profile), and immunoprecipitation from S35-labeled HeLa cell lysates (S35 IP). Comparators were included in the dermatomyositis (DM) group if they had autoantibodies recognizing Mi-2, NXP-2, TIF1-γ, or MDA-5. Alternatively, patients were classified as having Antisynthetase Syndrome (AS) if they had autoantibodies against Jo-1, PL-7, or PL-12. Patients were included in the Immune-mediated Necrotizing Myopathy (IMNM) group if they tested positive for anti-SRP or anti-HMGCR autoantibodies.^26^

### Investigations

All clinical assessments were performed by either a JH Myositis Center physician or another neuromuscular specialist at The Johns Hopkins University. Disease onset was defined as the time of onset of the initial symptom (presenting symptom) leading to a diagnosis of IBM. Date of diagnosis was defined as the date the diagnosis of IBM was made by a treating physician. Dates were derived as patient-reported or as otherwise noted in the medical record. If a specific date (MM/DD/YYYY) of onset or diagnosis was not available, the midpoint of the specified month or year was selected for the purposes of statistical analysis. Time to diagnosis was defined as the time interval between disease onset and IBM diagnosis date. Disease duration to first visit was defined as the time interval between disease onset and first visit to the JH Myositis Center. Initial symptoms and initial diagnoses other than IBM were derived as patient-reported or as otherwise noted in the medical record.

Manual muscle testing was recorded using the Medical Research Council (MRC) scale and converted to a ten-point Kendall scale for analysis. Manual muscle testing from the first JH Myositis Center visit was utilized for analysis in this study. The presence of dysphagia was assessed throughout the study based upon patient report of symptoms. Sera from a subset of IBM patients were tested for anti-NT5C1a antibodies by ELISA (ELISA MUP-44 [NT5C1A] Catalog #1675-4801G). Only electrodiagnostic studies performed at JH by an experienced electromyographer were included in this study. Conventional techniques were employed and JH electrodiagnostic laboratory normal values were utilized for analysis. The presence of peripheral neuropathy was defined as a sural sensory nerve action potential (SNAP) amplitude of < 9 μV. Muscle biopsy reports from both Johns Hopkins and outside institutions were included. Reports were reviewed by JH Myositis Center physicians and considered sufficient for diagnostic purposes if cardinal IBM histopathologic features were assessed. A subset of reports containing more detailed histopathologic descriptions was included for further analysis.

### Statistics

Dichotomous variables were expressed as percentages and absolute frequencies, and continuous features were reported as means and standard deviations (SD). Pairwise comparisons for categorical variables between groups were made using the chi-square test or Fisher’s exact test, as appropriate. The Student’s t-test was used to compare continuous variables among groups. CK, a highly positively skewed variable, was expressed as median, first, and third quartile for descriptive purposes. Linear and logistic regressions were used to adjust for potential confounding variables. Indirect standardization was used to compare mortality in IBM patients compared to the general population as described elsewhere.^27^

To account for the different number of visits per patient, the evolution of CK levels and muscle strength was studied using multilevel linear regression models with random slopes and random intercepts. Non-modifiable risk factors (sex, race, disease duration, and age at the onset of first symptoms) were used as adjusting covariates for the multivariate analysis. Generalized additive models (GAM) were applied to graphically analyze the longitudinal changes in strength and CK levels. All statistical analyses were performed using Stata/MP 14.1, and R v.4.0.3. A 2-sided p-value of 0.05 or less was considered significant with no correction for multiple comparisons.

### Standard Protocol Approvals, Registrations, and Patient Consents

This study was approved by an institutional review board at The Johns Hopkins Hospital. Written informed consent was obtain from all participants at the time of their enrollment in the JH Myositis Center Registry.

### Data availability

De-identified data, study protocol and statistical analysis plan will be made available in the case of a reasonable request providing that the proposed use of the data has been approved by an independent review committee and that investigators meet The Johns Hopkins University policies for data sharing and management.

## RESULTS

### Patient Characteristics

#### Clinical Criteria

Within the JH Myositis Center Registry, a total of 387 patients were identified with a listed diagnosis of IBM. Of these, 335 met at least Griggs possible, ENMC probable, or DDC criteria for IBM and were included in the subsequent analysis. Only a small percentage of IBM patients met either Griggs definite (2%) or ENMC clinico-pathologically defined (1%) criteria (Supplementary Table 1). This was primarily due to the lack of electron microscopy and evaluation for protein accumulations other than amyloid in routine muscle biopsy assessments. Nearly half of patients (46%) met criteria for Griggs, ENMC, and DDC, with 16% meeting only ENMC criteria, 15% meeting only DDC criteria, and 16% meeting both ENMC and DDC criteria (Figure 1).

**Figure 1.**
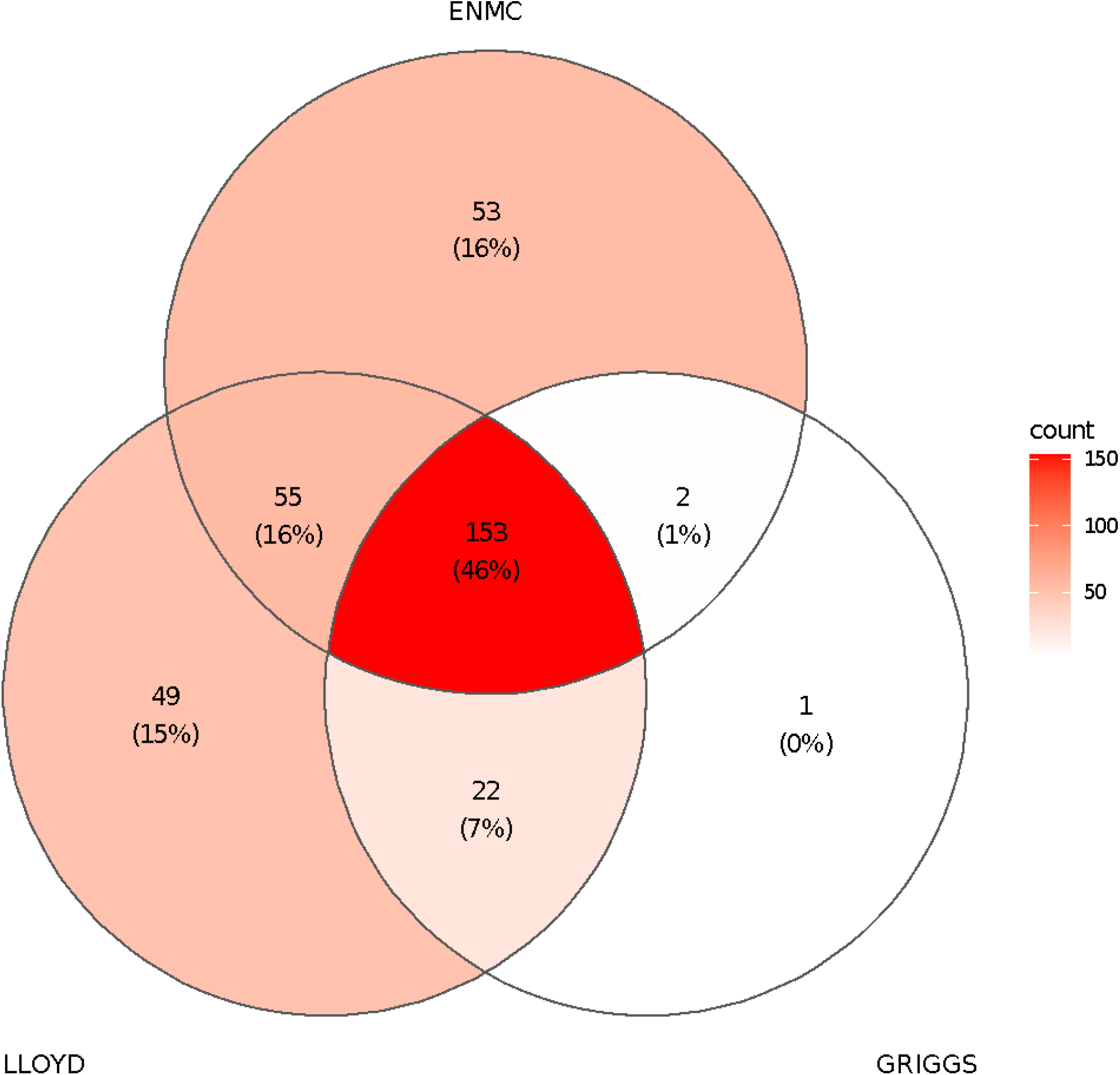
Overlap of diagnostic criteria in a large IBM cohort. ENMC: European Neuromuscular Centre criteria; GRIGGS: Griggs criteria; LLOYD: Data-Derived Criteria (also referred to in the literature as Lloyd-Greenberg criteria).

An additional 524 patients with other forms of autoimmune myositis were identified as a comparator group. These included 165 patients with IMNM, 199 patients with DM, and 160 patients with AS.

### IBM Clinical Characteristics

Baseline clinical information for the overall cohort are summarized in Supplementary Table 2. Compared with other types of myositis, IBM was more common in male (64%, p<0.001), White (84%, p<0.001), and older patients (median age at onset 58.7, p<0.001). The average time to diagnosis for IBM was longer than for other myositis groups (5.2 years, p<0.001). Only 4% of IBM patients reported dysphagia as a presenting symptom, which was less than IMNM (19%, p<0.001) or DM (19%, p<0.001). However, 60% developed symptoms of dysphagia during the course of their illness, which was significantly higher compared to patients with IMNM (45%, p=0.007) or the antisynthetase syndrome (38%, p<0.001) but not dermatomyositis (53%, p=0.99). A considerable number of patients (42%) were treated with immunosuppressive agents, although this was lower than any other type of myositis (p<0.001) in our study. Corticosteroids and methotrexate were the most common agents utilized. Compared with the other clinical groups, the overall mortality rate was higher in IBM than in IMNM; however, there were no significant differences in mortality between IBM and the general population (SMR 1.15, 95%CI 0.71-1.75).

Compared to patients with other types of myositis, those with IBM had more severe finger flexor and knee extensor weakness (Supplementary Table 3), showing a steady decline (about 0.33 Kendall scale strength-points per year) in these two muscle groups during the evolution of their disease (Figure 2). In contrast, hip flexors and arm abductors tended to be stronger in IBM than in IMNM during the first 10 years after the onset of the disease (Figure 2).

**Figure 2.**
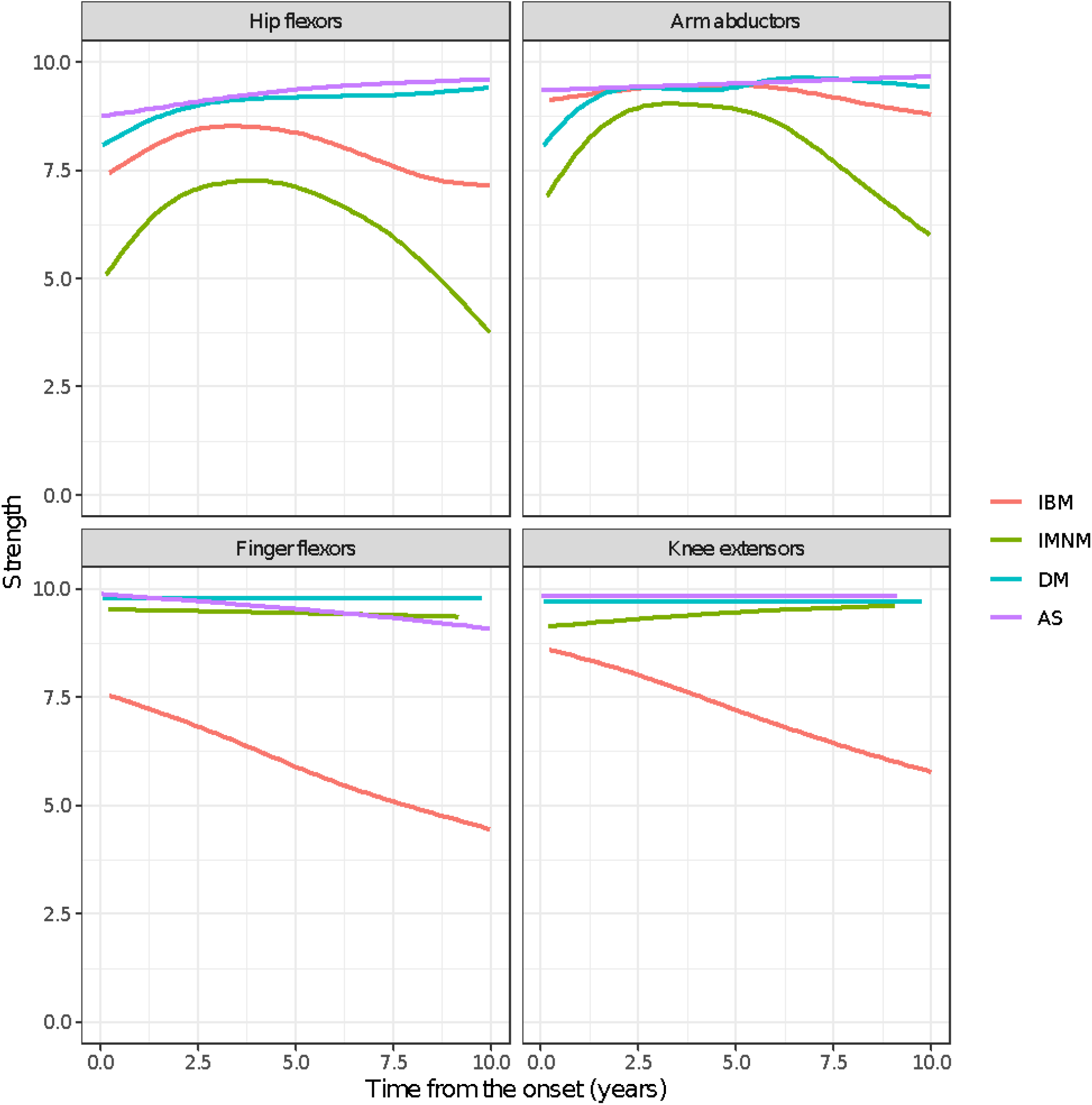
Evolution of strength in inclusion body myositis compared to other types of autoimmune myositis using generalized additive models.

Over half of IBM patients received an initial diagnosis other than IBM (Supplementary Table 4). The most common misdiagnoses were polymyositis (22%) and unspecified myopathy (13%). Patients with initial misdiagnoses were more likely to be female and to have a younger age at disease onset. Finger flexor weakness and rimmed vacuoles on muscle biopsy were less common in this group. Those with an initial diagnosis of polymyositis were more likely to be ANA positive compared to those initially diagnosed with IBM (50% versus 31%, p=0.04).

### IBM Ancillary Testing

The average duration of disease at the time of muscle biopsy was 5.5 years. The combination of endomysial inflammation, invasion of non-necrotic fibers, and rimmed vacuoles was seen in 43% of biopsies. An additional 41% contained at least two of these cardinal features, and 34% did not show rimmed vacuoles (Figure 3). The quadriceps muscle was the most common muscle selected for biopsy. The location of the muscle biopsy did not influence the number of pathologic features present, except for the more common presence of COX-negative fibers in the biceps (82%) or deltoid (83%) muscles compared to the quadriceps (47%).

**Figure 3.**
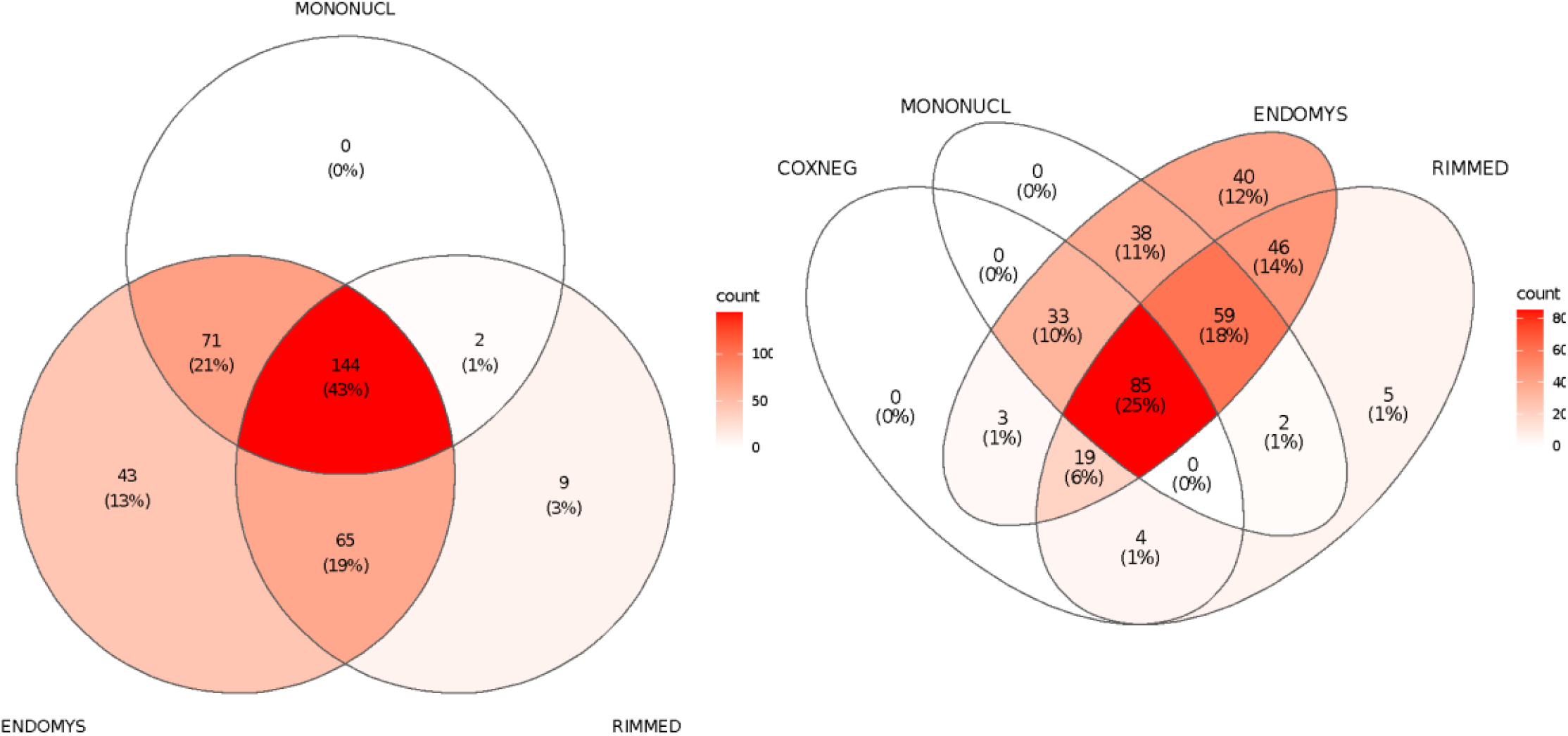
Cardinal IBM muscle biopsy features. **ENDOMYS:** Endomysial inflammation. **MONONUCL:** Mononuclear cell invasion of non-necrotic myocytes. **RIMMED:** Rimmed vacuoles. **COXNEG**: Cox-negative fibers.

Electrodiagnostic data was available for 285 (85%) IBM patients (Supplementary Table 5). Myopathic motor unit action potentials (small duration and small amplitude) were seen in nearly all patients with 44% also showing neurogenic units (large duration and large amplitude). Abnormal sural SNAP amplitudes (< 9 µV) were present in 46% of IBM patients compared to 24% of patients with other forms of myositis. However, multivariate analysis suggested that this difference may be accounted for by the older age of IBM patients at the time of EMG. The median CK in our IBM cohort was 426 IU/L with a Q1-Q3 range of 230-707 IU/L. A positive ANA was found in 40% of patients tested (n=199). Testing for the anti-NT5c1A autoantibody was performed by ELISA for 320 IBM patients. Forty-seven percent were positive for this autoantibody.

### IBM Heterogeneity

#### Race

Compared to other races, Black IBM patients were younger at the time of their first JH Myositis Center visit although this did not reach statistical significance. Black patients had significantly weaker arm abductors, hip flexors, and knee flexors (Table 1) at the first JH Myositis Center visit. They were less likely to report dysphagia during their disease course (45%) compared to non-Black patients (61%, p = 0.05). Black patients were more likely to have been misdiagnosed with polymyositis and treated with immunosuppressive agents. The median CK was higher in this group (676 IU/L, Q1-Q3 range 496-1035 IU/L) and neuropathic motor units were less commonly observed in electrodiagnostic studies (20%). Anti-NT5c1A autoantibodies were more commonly detected among Black patients (68%, p=0.02). There was no difference in muscle biopsy features across races.

**Table 1.**
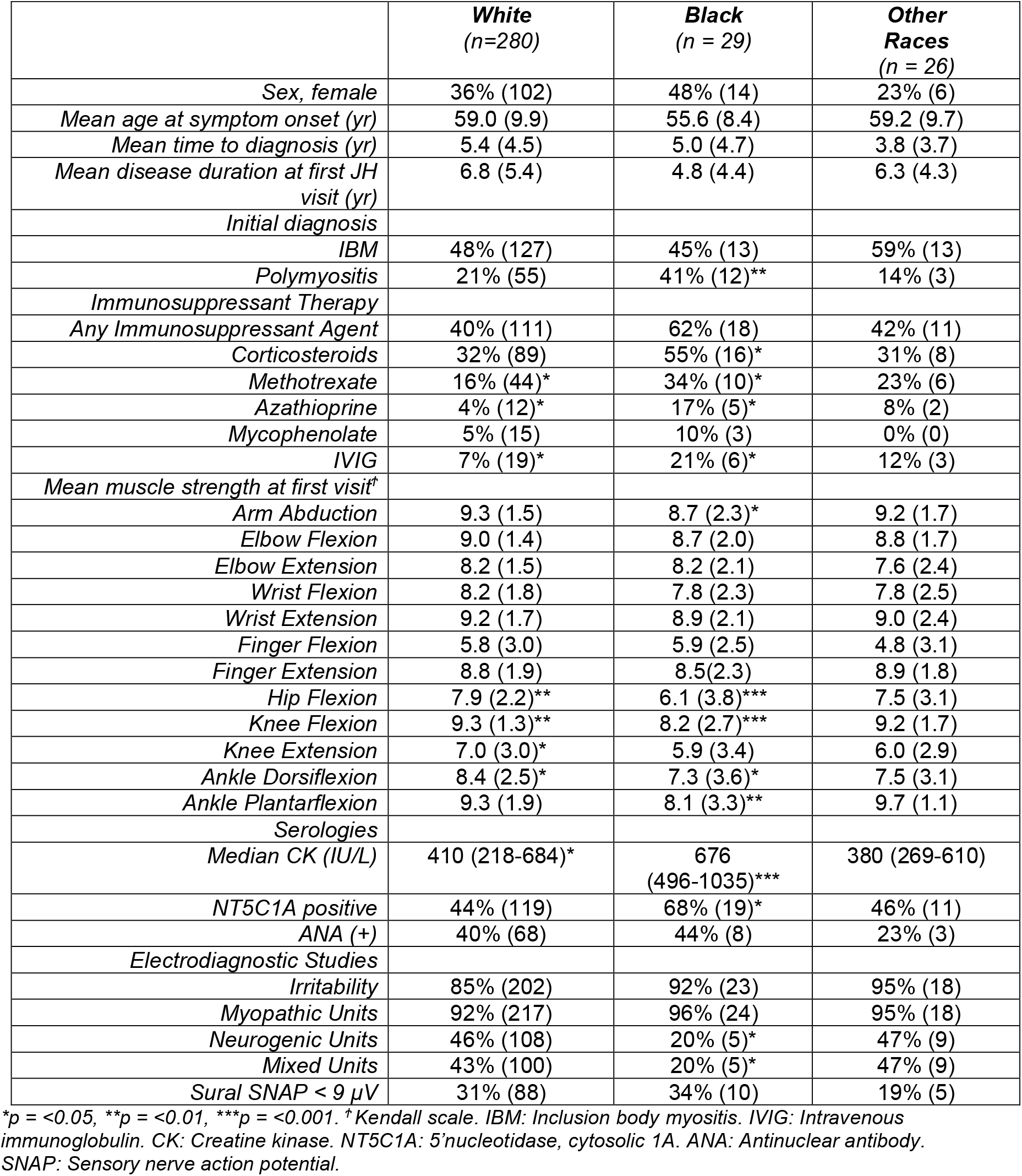
IBM characteristics by race grouping.

Multilevel regression models (independent of all the above-mentioned potential confounding factors) confirmed that Black patients had more severe weakness of hip flexors (−2.1 strength-points, p<0.001), arm abductors (−0.6 strength-points, p=0.02), and knee extensors (−1.3 strength-points, p=0.02), and had a trend toward weaker finger flexor muscles (−1.1 strength-points, p=0.06). It also confirmed that Black IBM patients have higher CK levels (p=0.001). Logistic regression showed that Black patients had lower odds of developing dysphagia (OR 0.4, p=0.05).

### Female Sex

Women had a longer time to diagnosis and were more likely to have been misdiagnosed with polymyositis than men (Table 2). Corticosteroid use was more common in women, although overall use of immunosuppressants did not differ between the sexes. Strength profiles showed that women had stronger knee extensors (p=0.003) and a trend towards having stronger finger flexors (p=0.06) (Figure 4). Women were more likely to develop dysphagia during their disease course (69%) compared to their male counterparts (55%, p = 0.02). Myopathic units were equally common between the sexes, although women tended to have less abnormal spontaneous activity on EMG.

**Table 2:** IBM characteristics by sex grouping.

|  | **Male**<br>(n = 213) | **Female**<br>(n = 122) |
| --- | --- | --- |
| *Mean age at symptom onset (yr)* | 59.2 (10.1) | 58.0 (9.1) |
| *Mean time to diagnosis (yr)* | 4.7 (4.2)** | 6.2 (4.8)** |
| *Mean disease duration at first JH visit (yr)* | 6.2 (5.2) | 7.3 (5.4) |
| *Initial diagnosis* |  |  |
| IBM | 55% (110)** | 37% (43)** |
| Polymyositis | 17% (34)** | 31% (36)** |
| *Immunosuppressant Therapy* |  |  |
| *Any Immunosuppressant Agent* |  |  |
| Corticosteroids | 30% (63)* | 41% (50)* |
| Methotrexate | 15% (33) | 22% (27) |
| Azathioprine | 6% (12) | 6% (7) |
| Mycophenolate | 4% (9) | 7% (9) |
| IVIg | 8% (17) | 9% (11) |
| *Mean muscle strength at first visit<sup>†</sup>* |  |  |
| Arm Abduction | 9.3 (1.7) | 9.2 (1.4) |
| Elbow Flexion | 9.0 (1.5) | 8.8 (1.5) |
| Elbow Extension | 8.1 (1.7) | 8.2 (1.6) |
| Wrist Flexion | 8.3 (1.7)* | 7.8 (2.1)* |
| Wrist Extension | 9.2 (1.8) | 9.0 (1.7) |
| Finger Flexion | 5.5 (3.1) | 6.1 (2.7) |
| Finger Extension | 8.9 (1.9) | 8.8 (1.9) |
| Hip Flexion | 7.7 (2.5) | 7.7 (2.4) |
| Knee Flexion | 9.2 (1.6) | 9.2 (1.4) |
| Knee Extension | 6.6 (3.2)* | 7.4 (2.6)* |
| Ankle Dorsiflexion | 8.1 (2.6) | 8.4 (2.7) |
| Ankle Plantarflexion | 9.3 (2.0) | 9.1 (2.2) |
| *Serologies* |  |  |
| Median CK (IU/L) | 467<br>(281-770)*** | 326<br>(174-522)*** |
| NT5C1A positive | 48% (96) | 45% (53) |
| ANA (+) | 34% (41) | 48% (38) |
| *Electrodiagnostic Studies* |  |  |
| Irritability | 92% (166)*** | 76% (77)*** |
| Myopathic Units | 94% (169) | 90% (90) |
| Neurogenic Units | 47% (83) | 39% (39) |
| Mixed Units | 44% (78) | 36% (36) |
| Sural SNAP < 9 μV | 32% (69) | 28% (34) |

\*p = <0.05, \*\*p = <0.01, \*\*\*p = <0.001. <sup>†</sup> Kendall scale. IBM: Inclusion body myositis. IVIg: Intravenous immunoglobulin. CK: Creatine kinase. NT5C1A: 5'nucleotidase, cytosolic 1A. ANA: Antinuclear antibody. SNAP: Sensory nerve action potential.

**Figure 4.**
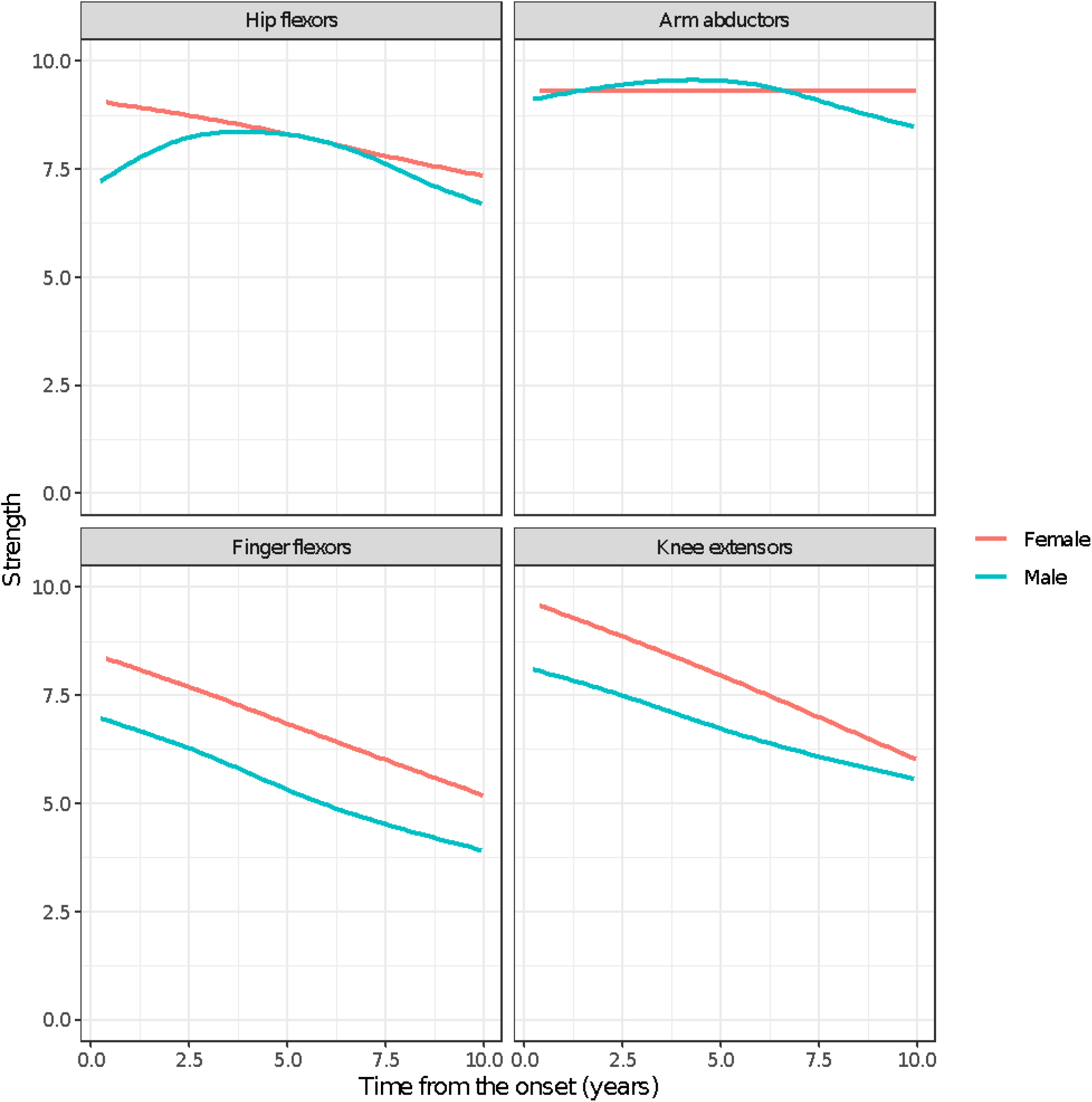
Evolution of strength in inclusion body myositis according to sex using generalized additive models.

Multilevel regression models confirmed that women have stronger finger flexors (0.8 strength-points, p=0.02) and knee extensor muscles (0.9 strength-points, p=0.009) with no significant difference in hip flexor or arm abductor strength (both p>0.05). CK was lower in females (p=0.02). Logistic regression showed that females develop dysphagia more frequently than males (OR 1.8, p=0.02).

### Age at Onset

Patients with a younger age at onset had a longer time to diagnosis and time to referral to the JH Myositis Center. Those with onset at < 50 years were also more likely to be initially misdiagnosed with polymyositis and treated with immunosuppressive therapy, especially methotrexate, mycophenolate, and IVIG. Muscle biopsy features did not differ between the age groups, although patients with a younger age at onset had a significantly longer disease duration at the time of biopsy, 7.3 years for the < 50 age group versus 2.9 years for the > 70 age group.

On univariate analysis, younger-onset patients had a more severe and diffuse pattern of weakness at the first visit, including more profound proximal weakness (Table 3). However, multivariate regression models demonstrated that this difference was driven by the longer disease duration at the first visit in the younger-onset age group. Older age at onset was associated with lower median CK levels (p<0.001). Logistic regression showed that there was no association between the prevalence of dysphagia and the age at onset, but there was a strong association between older age at onset and abnormal sural SNAP amplitudes (p<0.001).

**Table 3.** IBM characteristics by age of onset grouping.

|  | **< 50 yr**<br>(n = 66) | **50-61 yr**<br>(n = 117) | **61-70 yr**<br>(n = 102) | **>70 yr**<br>(n = 50) |
| --- | --- | --- | --- | --- |
| Sex, male | 64% (42) | 56% (65)* | 69% (70) | 72% (36) |
| Mean age at symptom onset (yr) | 44.8 (4.9)*** | 55.7 (2.8)*** | 64.3 (2.8)*** | 74.0 (3.3)*** |
| Mean time to diagnosis (yr) | 7.5 (5.7)*** | 5.8 (4.6) | 4.4 (3.3)* | 2.5 (1.9)*** |
| Mean disease duration at first visit (yr) | 9.2 (7.1)*** | 7.4 (5.2) | 5.6 (3.5)* | 2.9 (2.0)*** |
| Initial Diagnosis |  |  |  |  |
| IBM | 34% (22)** | 51% (54) | 52% (51) | 55% (26) |
| Polymyositis | 34% (22)* | 22% (23) | 20% (20) | 11% (5)* |
| Immunosuppressant Therapy |  |  |  |  |
| Any Immunosuppressant Agent | 55% (36)* | 41% (48) | 37% (38) | 36% (18) |
| Corticosteroids | 44% (29) | 31% (36) | 31% (32) | 32% (16) |
| Methotrexate | 27% (18)* | 17% (20) | 14% (14) | 16% (8) |
| Azathioprine | 11% (7) | 6% (7) | 4% (4) | 2% (1) |
| Mycophenolate | 14% (9)** | 4% (5) | 4% (4) | 0% (0) |
| IVIg | 15% (10)* | 12% (14) | 4% (4) | 0% (0) |
| Mean muscle strength at first visit† |  |  |  |  |
| Arm abduction | 8.5 (2.5)*** | 9.3 (1.4) | 9.5 (1.0) | 9.6 (0.7) |
| Elbow Flexion | 8.2 (2.2)*** | 9.0 (1.3) | 9.1 (1.2) | 9.4 (1.7)* |
| Elbow Extension | 7.6 (2.2)** | 8.2 (1.5) | 8.2 (1.6) | 8.6 (1.2)* |
| Wrist Flexion | 7.7 (2.2)* | 8.1 (2.0) | 8.2 (1.7) | 8.8 (1.2)* |
| Wrist Extension | 8.5 (2.5)*** | 9.2 (1.6) | 9.2 (1.7) | 9.8 (0.7)** |
| Finger Flexion | 5.1 (3.0) | 5.7 (3.2) | 5.9 (2.7) | 6.2 (3.0) |
| Finger Extension | 8.2 (2.5)** | 8.8 (1.9) | 8.9 (1.8) | 9.5 (0.8)* |
| Hip Flexion | 6.8 (3.3)*** | 7.9 (2.5) | 7.9 (2.2) | 8.1 (1.5) |
| Knee Flexion | 9.0 (2.0) | 9.2 (1.5) | 9.2 (1.5) | 9.6 (0.7)* |
| Knee Extension | 6.1 (3.5)* | 7.0 (3.0) | 6.9 (2.8) | 7.6 (2.6) |
| Ankle Dorsiflexion | 7.3 (3.5)** | 8.2 (2.7) | 8.6 (2.3) | 8.7 (1.5) |
| Ankle Plantarflexion | 9.0 (2.6) | 9.4 (1.8) | 9.1 (2.2) | 9.6 (1.3) |
| Serologies |  |  |  |  |
| Median CK (IU/L) | 460<br>(277-810) | 438<br>(249-720) | 430<br>(227-634) | 294<br>(167-477)** |
| NT5C1A positive | 49% (31) | 49% (56) | 41% (39) | 48% (23) |
| ANA (+) | 45% (17) | 34% (22) | 34% (23) | 57% (17)* |
| Electrodiagnostic Studies |  |  |  |  |
| Irritability | 89% (49) | 88% (85) | 79% (69)* | 95% (40) |
| Myopathic Units | 93% (51) | 95% (91) | 90% (77) | 95% (40) |
| Neurogenic Units | 29% (16)* | 43% (41) | 48% (41) | 57% (24) |
| Mixed Units | 27% (15)* | 41% (39) | 44% (37) | 55% (23)* |
| Sural SNAP < 9 µV | 23% (15) | 26% (31) | 32% (33) | 48% (24)** |

\*p = <0.05, \*\*p = <0.01, \*\*\*p = <0.001. † Kendall scale. IBM: Inclusion body myositis. IVIg: Intravenous immunoglobulin. CK: Creatine kinase. NT5C1A: 5'nucleotidase, cytosolic 1A. ANA: Antinuclear antibody. SNAP: Sensory nerve action potential.

### NT5C1A Status

Anti-NT5C1A-positive patients had higher median CK levels, but antibody status did not otherwise influence clinical characteristics, the pattern of weakness, muscle biopsy features, or electrodiagnostic findings.

## DISCUSSION

This study, including 335 IBM patients, is the largest cohort study of sporadic IBM to date. The general characteristics of our IBM cohort are similar to those previously reported, indicating that our findings are generalizable to the IBM population at large. Importantly, however, our large cohort also included significant numbers of female and Black patients, as well as patients with an age at onset < 50 years. These subpopulations have not been well described in the IBM literature to date, and few prior cohort studies have assessed the clinical phenotype in these IBM patient subpopulations. Although both Black and female patients demonstrated an overall pattern of weakness consistent with that typically seen in IBM, Black patients had significantly weaker proximal muscles and female patients had less prominent knee extension and finger flexion weakness. These characteristics may explain why both patient populations were more likely to be initially misdiagnosed with polymyositis and to be treated with corticosteroids.

Patients with an age of symptom onset < 50 years had more diffuse and severe weakness at the initial JHH visit compared to their older onset counterparts. However, this effect was confounded by a longer disease duration at the time of the first visit in the younger-onset group. Adjusting by this factor, we found that IBM patients with onset at an older age had more severe weakness in finger flexors, while other muscles showed similar levels of weakness. Other studies have reported more rapid progression in patients with an older age of onset, though this was not observed in our cohort.^9–11,14^

These findings suggest that Black and female patients may represent clinically distinct subgroups within IBM with unique disease trajectories and, potentially, different responses to therapeutic interventions. Although a distinct pattern was not confirmed in patients with a younger age at onset, our analysis was limited by constraining clinical assessments to visits at the JH Myositis Center. Thus, it is possible that this population did have a unique pattern of disease at onset which may be less prominent with more advanced disease.

In accordance with prior studies, a large percentage of our patients experienced a prolonged time to diagnosis, during which period many were initially misdiagnosed with polymyositis and treated with ineffective immunosuppressive therapies. We suspect that over-reliance on muscle biopsy features and under-reliance on physical exam (e.g. presence of finger flexor weakness and quadriceps>hip flexor weakness patterns) are major factors in the long delay to diagnosis that is common for IBM patients. Only 43% of muscle biopsies in our study contained all three cardinal features of IBM (endomysial inflammation, mononuclear cell invasion, and rimmed vacuoles). In particular, rimmed vacuoles were less common in our study (66%) compared to prior studies (75-100%).^8,11,14^ This likely reflects the increased reliance of Griggs and ENMC criteria for inclusion in prior published reports, both of which required the presence of rimmed vacuoles to meet definite or clinicopathologically defined IBM, respectively. This highlights the need for more accurate and less invasive diagnostics to identify IBM patients at earlier stages of disease when they may be more responsive to therapeutics.

Many of our patients also had electrodiagnostic evidence of peripheral neuropathy as defined by decreased sural SNAP amplitudes. Although a few small studies have reported evidence of peripheral nerve dysfunction in up to 30% of IBM patients, the involvement of peripheral nerves has not been extensively studied.^28,29^ In the current study, we found evidence of axonal loss in 46% of IBM patients, well beyond the reported estimates of peripheral neuropathy in the general population (2-8%).^30^ However, when compared with patients with other forms of autoimmune myositis, the prevalence of sural SNAP abnormalities was found to be driven by the older age at EMG/NCS rather than by the presence of IBM. Detailed studies of electrodiagnostic data in IBM are needed to explore this further.

Our study has several limitations. Data from clinical assessments were constrained to visits at the JH Myositis Center at which point patients had had symptoms for 6.6 years on average. Although female and Black IBM patients did have different patterns of weakness by this time, their initial symptoms may have differed more dramatically. In addition, younger-onset patients may have had a distinct phenotype at presentation which became obscured by more advanced disease.

In conclusion, sporadic IBM has long been considered a disease predominately affecting older White men. Our cohort included a significant number of female, Black, and younger-onset patients and demonstrated a unique clinical profile in both female and Black patients that may explain the propensity for initial misdiagnosis and ineffective immunosuppressant agent use in these populations. Further longitudinal studies are needed to confirm and further characterize these findings; however, it is important to recognize the presence of these subgroups as they may have different responses to therapies, and their presence may influence the design of future clinical trials in IBM. More sensitive and accessible diagnostics are needed to identify IBM patients at earlier stages and within these less well-recognized subpopulations to further our understanding of clinical heterogeneity within this disorder.

## Acknowledgements

This study was funded by The Sandra and Malcolm Berman Brain & Spine Institute, The Peter and Carmen Lucia Buck Foundation, and the Intramural Research Program of the National Institute of Arthritis and Musculoskeletal and Skin Diseases, National Institutes of Health. We thank these individuals and institutions for their contribution.

---

**Supplementary Table 1.** Patients meeting IBM diagnostic criteria.

| ***Diagnostic Criteria*** | ***% (n)*** |
| --- | --- |
| ***Griggs*** |  |
| *Definite* | 2% (7) |
| *Possible* | 53% (177) |
| ***ENMC 2011*** |  |
| *Clinico-Pathologically Defined* | 1% (2) |
| *Clinically Defined* | 53% (177) |
| *Probable* | 78% (261) |
| ***Lloyd-Greenberg*** | 83% (279) |

*IBM: Inclusion body myositis*

**Supplementary Table 2.** Overall clinical characteristics of inclusion body myositis compared to other types of autoimmune myositis.

|  | IBM | IMNM | Multivariate<br>p-value | DM | Multivariate<br>p-value | AS | Multivariate<br>p-value |
| --- | --- | --- | --- | --- | --- | --- | --- |
|  | (n=335) | (n=165) |  | (n=199) |  | (n=160) |  |
| **Female sex** | 36% (122) | 62% (103)*** | .. | 73% (145)*** | .. | 72% (115)*** | .. |
| **Race** |  |  |  |  |  |  |  |
| **White** | 84% (280) | 68% (112)*** | .. | 75% (149)* | .. | 58% (93)*** | .. |
| **Black** | 9% (29) | 24% (40)*** | .. | 14% (27) | .. | 31% (50)*** | .. |
| **Other races** | 8% (26) | 8% (13) | .. | 12% (23) | .. | 11% (17) | .. |
| **Age at onset (years)** | 58.7 (9.8) | 51.1 (14.6)*** | .. | 47.8 (14.9)*** | .. | 46.1 (12.8)*** | .. |
| **Time to diagnosis** | 5.2 (4.5) | 1.8 (4.5)*** | < 0.001 | 1.1 (2.0)*** | < 0.001 | 1.1 (1.6)*** | < 0.001 |
| **Time of follow-up (years)** | 2.3 (2.8) | 3.9 (3.7)*** | < 0.001 | 4.3 (3.6)*** | < 0.001 | 4.6 (3.9)*** | < 0.001 |
| **Time to first JH visit (years)** | 6.6 (5.3) | 2.4 (3.2)*** | < 0.001 | 1.5 (3.1)*** | < 0.001 | 2.2 (2.9)*** | < 0.001 |
| **Number of visits per participant** | 4.2 (4.7) | 9.1 (8.9)*** | < 0.001 | 9.6 (7.0)*** | < 0.001 | 9.7 (6.8)*** | < 0.001 |
| **Death during follow-up** | 7% (25) | 3% (5)* | 0.6 | 5% (9) | 0.9 | 9% (14) | 0.09 |
| **Dysphagia at onset** | 4% (12) | 19% (31)*** | < 0.001 | 19% (38)*** | < 0.001 | 6% (10) | 0.5 |
| **Dysphagia during follow-up** | 60% (201) | 45% (74)** | 0.007 | 53% (106) | 1.0 | 38% (60)*** | < 0.001 |
| **Treatments** |  |  |  |  |  |  |  |
| **Any immunosuppressant** | 42% (140) | 87% (143)*** | < 0.001 | 96% (192)*** | < 0.001 | 97% (155)*** | < 0.001 |
| **Corticosteroids** | 34% (113) | 74% (122)*** | < 0.001 | 83% (166)*** | < 0.001 | 94% (151)*** | < 0.001 |
| **Azathioprine** | 6% (19) | 26% (43)*** | < 0.001 | 28% (56)*** | < 0.001 | 52% (83)*** | < 0.001 |
| **Methotrexate** | 18% (60) | 52% (86)*** | < 0.001 | 53% (105)*** | < 0.001 | 42% (67)*** | 0.005 |
| **Mycophenolate** | 5% (18) | 19% (31)*** | 0.003 | 36% (72)*** | < 0.001 | 41% (65)*** | < 0.001 |
| **IVIG** | 8% (28) | 42% (69)*** | < 0.001 | 47% (94)*** | < 0.001 | 36% (57)*** | < 0.001 |
| **Rituximab** | 0% (0) | 24% (40)*** | .. | 17% (33)*** | .. | 24% (38)*** | .. |

*Dichotomous variables were expressed as a percentage (count) and continuous variables as mean (SD). Univariate comparisons of continuous variables were made using Student's t-test while dichotomous variables were compared either using the chi-squared test or Fisher's exact test, as appropriate. Multivariate comparisons were performed using logistic regression for dichotomous variables. All multivariate comparisons were adjusted by age at onset, sex, race, and duration of follow-up (time from the onset to the last follow-up). IBM: Inclusion body myositis. IMNM: Immune medication necrotizing myositis. DM: Dermatomyositis. AS: Anti-synthetase syndrome. IVIG: Intravenous immunoglobulin.*

**Supplementary Table 3.** Weakness pattern in inclusion body myositis compared to other types of autoimmune myositis at the first JH visit.

|  | **IBM** | **IMNM** | **Multivariate**<br>**p-value** | **DM** | **Multivariate**<br>**p-value** | **AS** | **Multivariate**<br>**p-value** |
| --- | --- | --- | --- | --- | --- | --- | --- |
|  | **(n=335)** | **(n=165)** |  | **(n=199)** |  | **(n=160)** |  |
| **Major muscle regions** |  |  |  |  |  |  |  |
| **Proximal upper extremity weakness** | 84% (283) | 74% (122)** | < 0.001 | 59% (117)*** | < 0.001 | 39% (63)*** | < 0.001 |
| **Distal upper extremity weakness** | 97% (326) | 16% (27)*** | < 0.001 | 27% (54)*** | < 0.001 | 14% (22)*** | < 0.001 |
| **Proximal lower extremity weakness** | 93% (311) | 87% (143)* | 0.01 | 65% (129)*** | < 0.001 | 56% (90)*** | < 0.001 |
| **Distal lower extremity weakness** | 61% (203) | 7% (12)*** | < 0.001 | 12% (23)*** | < 0.001 | 2% (3)*** | < 0.001 |
| **Individual muscle group strength <sup>†</sup>** |  |  |  |  |  |  |  |
| **Arm abductors** | 9.2 (1.6) | 8.0 (2.0)*** | < 0.001 | 8.4 (2.4)*** | < 0.001 | 9.3 (1.4) | 0.2 |
| **Elbow flexors** | 8.9 (1.5) | 8.9 (1.7) | 0.4 | 9.1 (1.5) | 0.4 | 9.7 (0.6)*** | < 0.001 |
| **Elbow extensors** | 8.1 (1.7) | 8.7 (1.7)** | 0.01 | 8.9 (1.6)*** | 0.007 | 9.6 (0.9)*** | < 0.001 |
| **Wrist flexors** | 8.2 (1.9) | 9.8 (0.7)*** | < 0.001 | 9.7 (0.8)*** | < 0.001 | 9.9 (0.4)*** | < 0.001 |
| **Wrist extensors** | 9.1 (1.8) | 9.8 (0.6)*** | 0.008 | 9.7 (1.0)*** | 0.03 | 9.9 (0.4)*** | 0.004 |
| **Finger flexors** | 5.7 (3.0) | 9.7 (0.7)*** | < 0.001 | 9.8 (0.6)*** | < 0.001 | 9.8 (0.6)*** | < 0.001 |
| **Finger extensors** | 8.8 (1.9) | 9.9 (0.4)*** | < 0.001 | 9.7 (1.0)*** | < 0.001 | 9.8 (0.5)*** | 0.002 |
| **Hip flexors** | 7.7 (2.5) | 5.9 (3.1)*** | < 0.001 | 8.1 (2.5) | 0.8 | 8.8 (1.8)*** | < 0.001 |
| **Hip extensors** | 9.8 (0.9) | 8.9 (2.3)*** | 0.004 | 9.5 (1.5)* | 0.02 | 9.8 (0.7) | 0.2 |
| **Knee flexors** | 9.2 (1.5) | 9.0 (1.8) | 0.6 | 9.8 (0.9)*** | 0.005 | 9.9 (0.4)*** | 0.004 |
| **Knee extensors** | 6.9 (3.0) | 9.2 (1.7)*** | < 0.001 | 9.7 (1.0)*** | < 0.001 | 9.8 (0.5)*** | < 0.001 |
| **Ankle flexors** | 8.2 (2.6) | 9.8 (0.5)*** | < 0.001 | 9.8 (1.0)*** | < 0.001 | 9.9 (0.3)*** | < 0.001 |
| **Ankle extensors** | 9.3 (2.0) | 9.8 (0.5)* | 0.2 | 9.9 (0.5)*** | 0.03 | 10.0 (0.0)** | 0.07 |

Variables were expressed as a mean (SD). Univariate comparisons were compared using Student's *t*-test. Multivariate comparisons were performed using linear regression. All multivariate comparisons were adjusted by age at onset, sex, race, and duration of follow-up (time from the onset to the last follow-up). <sup>†</sup> Kendall scale. IBM: Inclusion body myositis. IMNM: Immune medication necrotizing myositis. DM: Dermatomyositis. AS: Anti-synthetase syndrome.

**Supplementary Table 4:**
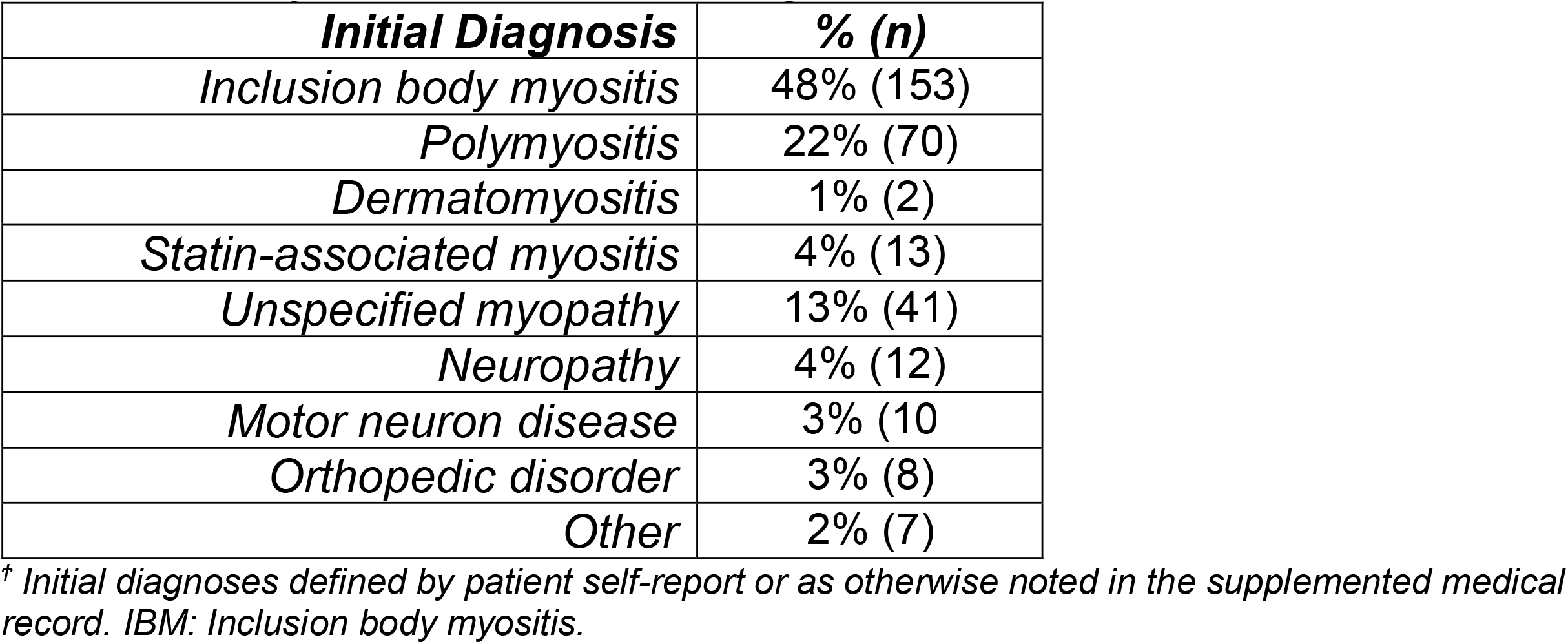
IBM initial diagnoses ^*Ϯ*^.

| ***Initial Diagnosis*** | ***% (n)*** |
| --- | --- |
| *Inclusion body myositis* | 48% (153) |
| *Polymyositis* | 22% (70) |
| *Dermatomyositis* | 1% (2) |
| *Statin-associated myositis* | 4% (13) |
| *Unspecified myopathy* | 13% (41) |
| *Neuropathy* | 4% (12) |
| *Motor neuron disease* | 3% (10) |
| *Orthopedic disorder* | 3% (8) |
| *Other* | 2% (7) |

<sup>†</sup> Initial diagnoses defined by patient self-report or as otherwise noted in the supplemented medical record. IBM: Inclusion body myositis.

**Supplementary Table 5.** IBM ancillary studies.

| ***Variable*** | ***Result***<br>***(n)*** |
| --- | --- |
| *Electrodiagnostic Studies* |  |
| *Irritability* | 86% (243) |
| *Myopathic units* | 93% (259) |
| *Neurogenic units* | 44% (122) |
| *Mixed units* | 41% (114) |
| *Sural SNAP < 9 μV* | 31% (103) |
| *Serologies* |  |
| *Median CK (IU/L)* | 426 (230-707) |
| *Anti-NT5C1A (+)* | 47% (149) |
| *ANA (+)* | 40% (79) |

IBM: Inclusion body myositis. SNAP: Sensory nerve action potential. CK: Creatine kinase. Anti-NT5C1A: Anti-5'nucleotidase, cytosolic 1A. ANA: Antinuclear antibody.

## References

1. Dalakas MC, Koffman B, Fujii M, Spector S, Sivakumar K, Cupler E. A controlled study of intravenous immunoglobulin combined with prednisone in the treatment of IBM. Neurology. 2001;56:323–327.

2. Badrising UA, Maat-Schieman MLC, Ferrari MD et al. Comparison of weakness progression in inclusion body myositis during treatment with methotrexate or placebo. Ann Neurol. 2002;51:369–372.

3. The Muscle Study Group. Randomized pilot trial of high-dose betaINF-1a in patients with inclusion body myositis. Neurology. 2004;63:718–720.

4. Barohn RJ, Herbelin L, Kissel JT, et al. Pilot trial of etanercept in the treatment of inclusion-body myositis. Neurology. 2006;66(suppl 1):S123–S124.

5. Dalakas MC, Rakocevic G, Schmidt J, et al. Effect of alemtuzumab (CAMPATH 1-H) in patients with inclusion-body myositis. Brain. 2009;132:1536–1544.

6. Benveniste O, Stenzel W, Hilton-Jones D, Sandri M, Boyer O, van Engelen BGM. Amyloid deposits and inflammatory infiltrates in sporadic inclusion body myositis: the inflammatory egg comes before the degenerative chicken. Acta Neuropathol. 2015;129:611–624.

7. Greenberg SA, Pinkus JL, Amato AA, Kristenson T, Dorfman DM. Association of inclusion body myositis with T cell large granular lymphocytic leukemia. Brain. 2016;139:1348–1360.

8. Lotz BP, Engel AG, Nishino H, Stevens JC, Litchy WJ. Inclusion body myositis: Observations in 40 patients. Brain. 1989;112:727–747.

9. Peng A, Koffman BM, Malley JD, Dalakas MC. Disease progression in sporadic inclusion body myositis: Observations in 78 patients. Neurology. 2000;55:296–298.

10. Badrising UA, Maat-Schieman MLC, Van Houwelingen JC, et al. Inclusion body myositis: Clinical features and clinical course of the disease in 64 patients. J Neurol. 2005;252:1448–1454.

11. Benveniste O, Guiguet M, Freebody J, et al. Long-term observational study of sporadic inclusion body myositis. Brain. 2011;134:3176–3184.

12. Cox FM, Titulaer MJ, Sont JK, Wintzen AR, Verschuuren JJGM, Badrising UA. A 12-year follow-up in sporadic inclusion body myositis: An end stage with major disabilities. Brain. 2011;134:3167–3175.

13. Cortese A, Machado P, Morrow J, et al. Longitudinal observational study of sporadic inclusion body myositis: implications for clinical trials. Neuromuscul Disord. 2013;23:404–412.

14. Hori H, Yamashita S, Tawara N, et al. Clinical features of Japanese patients with inclusion body myositis. J Neurol Sci. 2014;346:133–137.

15. Oldroyd AGS, Lilleker JB, Williams J, Chinoy H, Miller JAL. Long-term strength and functional status in inclusion body myositis and identification of trajectory subgroups. Muscle Nerve. 2020;62:76–82.

16. Shelly S, Mielke MM, Mandrekar J, et al. Epidemiology and natural history of inclusion body myositis. Neurology. 2021;96:e2653–e2661.

17. Naddaf E, Shelly S, Mandrekar J, et al. Survival and associated comorbidities in inclusion body myositis. Rheumatology. 2021;00:1–9.

18. Price MA, Barghout V, Benveniste O, et al. Mortality and causes of death in patients with sporadic inclusion body myositis: survey study based on the clinical experience of specialists in Australia, Europe and the USA. J Neuromuscula. Dis 2016;3:67–75.

19. Sangha G, Yao B, Lunn D, et al. Longitudinal observational study investigating outcome measures for clinical trials in inclusion body myositis. J Neurol Neurosurg Psychiatry. 2021;92:854–862.

20. Lilleker J, Rietveld A, Pye S, et al. Cytosolic 5’-nucleotidase 1A autoantibody profile and clinical characteristics in inclusion body myositis. Ann Rheum Dis. 2017;76:862–868.

21. Ikenaga C, Findlay AR, Goyal NA, et al. Clinical utility of anti-cytosolic 5’-nucleotidase 1A antibody in idiopathic inflammatory myopathies. Ann Clin Transl Neurol. 2021;8:571–578.

22. Paul P, Liewluck T, Ernste FC, Mandrekar J, Milone M. Anti-cN1A antibodies do not correlate with specific clinical, electromyographic, or pathologic findings in sporadic inclusion body myositis. Muscle Nerve. 2021;63:490–496.

23. Griggs RC, Askanas V, DiMauro S, et al. Inclusion body myositis and myopathies. Ann Neurol. 1995;38:705–13.

24. Rose MR, ENMC IBM Working Group. 188^th^ ENMC International Workshop: Inclusion body myositis, 2-4 December 2011, Naarden, The Netherlands. Neuromuscul Disord. 2013;23:1044–1055.

25. Lloyd, TE, Mammen AL, Amato AA, Weiss MD, Needham M, Greenberg SA. Evaluation and construction of diagnostic criteria for inclusion body myositis. Neurology. 2014;83:426–433.

26. Casal-Dominguez M, Pinal-Fernandez I, Pak K, et al. Performance of the 2017 EULAR/ACR classification criteria for inflammatory myopathies in patients with myositis-specific autoantibodies. Arthritis Rheumatol. 2021. Doi:10.1002/art.41964.

27. Tiniakou E, Pinal-Fernandez I, Lloyd T, et al. More severe disease and slower recovery in younger patients with anti-3-hydroxy-3-methylglutaryl-coenzyme A reductase-associated autoimmune myopathy. Rheumatology. 2017;56:787–794.

28. Lee JH, Boland-Freitas R, Liang C, Howells J, Ng K. Neuropathy in sporadic inclusion body myositis: a multi-modality neurophysiological study. Clin Neurophysiol. 2020;121:2766–2776.

29. Joy JL, Oh SJ, Baysal AI. Electrophysiological spectrum of inclusion body myositis. Muscle Nerve. 1990;13:949–951.

30. Martyn CN, Hughes RAC. Epidemiology of peripheral neuropathy. J Neurol Neurosurg Psychiatry. 1997;62:310–318.

